# Parkinson’s Disease With and Without History of Agent Orange Exposure: A Prospective Study of US Veterans

**DOI:** 10.64898/2026.01.22.26344300

**Authors:** Vivikta Iyer, Cole Zweber, Ian Zurlo, Pedram Golnari, Katrina Prantzalos, Brenna M. Lobb, Dakota Clarke, James T. Boyd, Satya S. Sahoo, Curtis Tatsuoka, Deepak K. Gupta, Amie L. Hiller

## Abstract

Environmental neurotoxins have been implicated in the pathogenesis of Parkinsons disease (PD), with Agent Orange (AO) recognized as a presumptive service-connected exposure among U.S. Veterans. However, prospective data examining potential clinical differences associated with AO exposure remain limited.

We conducted a multicenter prospective cohort study of U.S. Veterans with PD to compare demographic and clinical characteristics between individuals with and without a history of AO exposure. Clinical assessments included cognitive testing, motor and non-motor symptom scales, activities of daily living, and a global composite outcome.

Among 40 Veterans analyzed, no statistically significant differences were observed between exposed and unexposed groups on univariate analyses. However, trend-level differences suggested lower cognitive performance and greater patient-reported motor disability among AO-exposed Veterans.

While exploratory and limited by sample size, these findings suggest potential clinically meaningful distinctions that warrant confirmation in larger cohorts. Improved characterization of PD phenotypes associated with environmental toxin exposure may inform prognostic counselling and care of affected Veterans.

## Manuscript

The increasing global epidemic of Parkinson’s disease (PD) highlights the need for a deeper understanding of its different presentations and underlying etiologies. U.S. Veterans represent an important subgroup of PD with respect to potential clinical and pathophysiological differences^1^. PD can be a service-connected condition for U.S. Veterans, with presumptive connection for those who served in specific locations or timeframes, and non-presumptive service connection for those whose symptoms began during active military service, within one year of leaving service, or due to a traumatic brain injury (TBI)^2^. These service connections are based on evidence supporting an elevated risk of PD in association with environmental toxins^3^ or TBI, both of which can occur as occupational exposures during military service.

Some neurotoxins, such as MPTP (1-methyl-4-phenyl-1,2,3,6-tetrahydropyridine), rotenone, paraquat, and Agent Orange (AO), have been shown to induce PD pathophysiology through various mechanisms, including mitochondrial dysfunction, generation of superoxide free radicals, and dopaminergic cell death^4^. Among these, AO is considered the most significantly associated with PD in the context of U.S. Veterans. AO is composed of 2,4-dichlorophenoxyacetic acid (2,4-D) and 2,4,5-trichlorophenoxyacetic acid (2,4,5-T), as well as dioxin (2,3,7,8-tetrachlorodibenzo-p-dioxin (TCDD). Increasing evidence indicates that exposure to AO is associated with neurologic disorders, particularly neurodegeneration and parkinsonism^5^. A history of AO exposure makes PD a presumptive service-connected disease for US Veterans and makes them eligible to receive health care and disability compensation through the US Department of Veterans Affairs (VA). Moreover, a retrospective clinical research study in Korean Veterans showed that PD patients with history of exposure to AO exhibited different clinical phenotypes compared to those without such history of exposure^6^. However, to the best of our knowledge, no prospective data are available on potential differences in PD with a history of AO exposure in US Veterans.

Our multicenter, prospective cohort study was conducted at the University of Vermont Medical Center (UVMMC), VA Portland Health Care System (VHAPORHCS), and Oregon Health Science University (OHSU). Our study was approved by the single IRB (WCG) and Office of Human Research Oversight of the DoD.

We recruited a total of 100 patients, 50 from each site. Two subjects were excluded because their AO exposure history was unknown. The distribution of the primary and secondary grouping variable in the remaining 98 PD patients was assessed. The primary grouping variable, history of AO exposure was 31.63% (31 out of 98) from the VHAPORHCS/OHSU site (n = 28), it was determined by asking the patient to recall if they had any exposure, which was complemented with VA records to ensure best possible accuracy. The secondary grouping variable, Veteran status 44.9% (44 out of 98) were Veterans, again with most being from the VHAPORHCS/OHSU site (n = 40), was determined based on the patient’s enrollment in VA healthcare, irrespective of their VA service connection for PD or any other conditions. Given the high proportion of subjects being from the VHAPORHCS/OHSU site, potential confounding of effect of site was eliminated by restricting subsequent data analysis to the cohort of Veteran PD patients from the VHAPORHCS/OHSU site conferring a final cohort of 40 patients.

Patient information across the categories of demographics, health profile, history, exam, and clinical scales was collected using our clinical decision support (CDS) tool for PD (CDS-PD)^7^. We assessed for significant differences between the two groups of primary grouping variable in terms of baseline demographics (age, education, sex) and clinical characteristics (disease duration, Movement Disorder Society-Sponsored Unified Parkinson’s Disease Rating Scale (MDS-UPDRS) scores, Montreal Cognitive Assessment (MoCA), University of Pennsylvania

Smell Identification Test (UPSIT), and Modified Schwab & England Activity of Daily Living (MSEADL). We then looked for potential prognostic differences between the two groups using the Global Composite Outcome (GCO)^8^, which is an aggregate numeric indicator which merges the most clinical domains of the MDS-UPDRS, MSEADL, and MoCA, such that higher GCO values indicate worse global disease burden. Of note, the GCO as a prognostic marker has been shown to be significantly different at baseline and in follow-up among ‘mild motor-predominant’, ‘diffuse malignant’, and ‘intermediate’ phenotypes of PD.

In the cohort chosen for analysis, the prevalence of history of AO exposure was 70% (28 out of 40). There were no significant differences in demographics or clinical characteristics, including GCO, between the two groups on univariate analysis utilizing Chi-square for categorical variables and Student’s t-test for continuous variables (p < 0.05; table 1). Given the exploratory nature of this study, p-values were not adjusted for multiple comparisons and results were used as hypothesis-generating. Specifically, we then calculated and compared estimated marginal means (EMM) of selected variables between the two groups after adjusting for covariates (age, sex, education, and disease duration; p < 0.10).

**Table 1:** Chi-square test for categorical and Student-t test for continuous variables in the exposed and unexposed groups.

| Variable (unit) | All Veterans<br>(n = 40) | Exposed<br>(n = 28) | Unexposed<br>(n = 12) | p-value |
| --- | --- | --- | --- | --- |
| Sex (male) | 39<br>(97.5%) | 28<br>(100%) | 11<br>(91.66%) | 0.658 |
| MCI*<br>#(n=39) | 26<br>(66.66%) | 21<br>(75%) | 5<br>(45.45%) | 0.166 |
| Positive Family History of PD in FDR* | 3<br>(7.5%) | 2<br>(7.14%) | 1<br>(8.33%) | 1.000 |
| Age<br>(years) | 76.58 ± 4.09<br>(69 - 88) | 77.07 ± 3.59<br>(69 - 88) | 75.42 ± 5.05<br>(69-86) | 0.319 |
| Education (years) | 15.06 ± 3.62<br>(0-20) | 14.82 ± 3.71<br>(0-20) | 15.50 ± 3.50<br>(8-20) | 0.587 |
| Disease Duration (years) | 5.30 ± 4.05<br>(1-20) | 5.79 ± 4.19<br>(1-14) | 4.17 ± 3.61<br>(1-20) | 0.228 |
| MoCA<br>#(n=39) | 23.18 ± 4.56<br>(7 - 30) | 22.36 ± 4.44<br>(7 - 29) | 25.27 ± 4.36<br>(17 - 30) | **0.077** |
| UPSIT | 16.76 ± 5.82<br>(8 - 30) | 17.15 ± 6.60<br>(8 - 30) | 15.82 ± 3.46<br>(11-21) | 0.427 |
| MSEADL | 72.25 ± 20.19<br>(20 - 100) | 71.07 ± 20.61<br>(20 - 90) | 75 ± 19.77<br>(30 - 100) | 0.575 |
| MDS-UPDRS Part I | 14.60 ± 7.24<br>(3 - 34) | 15.54 ± 8.00<br>(3 - 34) | 12.42 ± 4.58<br>(4 - 18) | 0.130 |
| MDS-UPDRS Part II | 17.95 ± 10.45<br>(4 - 47) | 19.75 ± 10.50<br>(4 - 47) | 13.75 ± 9.43<br>(4 -39) | **0.088** |
| MDS-UPDRS Part III | 42.70 ± 15.14<br>(16 - 85) | 43.71 ± 14.32<br>(23 - 85) | 40.33 ± 17.33<br>(16 - 67) | 0.560 |
| MDS-UPDRS Part IV | 3.95 ± 3.81<br>(0 - 11) | 3.79 ± 3.77<br>(0 - 11) | 4.33 ± 4.03<br>(0 -10) | 0.693 |
| GCO | 0.00 ± 1.73<br>(-3.80 to 3.46) | 0.13 ± 1.79<br>(-3.80 to 3.46) | -0.30 ± 1.61<br>(-3.15 to 2.06) | 0.459 |

Abbreviations: MCI = Mild Cognitive Impairment, MoCA = Montreal Cognitive Assessment, MDS-UPDRS = Movement Disorders Society Unified Parkinsons Disease Rating Scale, FDR = First Degree Relatives, PD = Parkinson's Disease, UPSIT = University of Pennsylvania Smell Identification Test, MSEADL = Modified Schwab and England Activities of Daily Living, TD = Tremor Dominant, PIGD = Postural Instability and Gait Disorder, GCO = Global Composite Outcome.
\* = Fisher's exact test was used where expected cell counts were <5.

Of note, there was a trend towards significant differences in two clinical characteristics: MoCA score, which represents cognitive function, was lower at 22.36 ± 4.44 in exposed versus 25.27 ± 4.36 in the unexposed group; MDS-UPDRS Part II score, which represents patient reported motor aspects of activities of daily living, was higher at 19.75 ± 10.50 in exposed versus 13.75 ± 9.43 in unexposed. To further estimate potential differences in these two variables, we calculated their EMM and found that the difference for the two groups was −2.92 (adjusted *p*-value of 0.072) and 6.0 (adjusted *p*-value 0.096) for the MoCA and the MDS-UPDRS Part II scores, respectively.

Our prospective clinical research study found no statistically significant differences in MoCA, MDS-UPDRS, MSEADL, and GCO, in US Veterans with and without AO exposure. However, trends in MoCA and MDS-UPDRS Part II suggest potential clinically meaningful distinctions that likely require a larger sample size for confirmation. While previous retrospective research in Korean Veterans ^21^ identified differences in objective motor signs (Part III), our findings highlight trends in measures of patient-reported motor experiences and cognitive function. These discrepancies may stem from differences in study design and the distinct genetic or lifestyle factors of US Veterans and may also indicate that PD phenotypes associated with history of AO exposure may preferentially reflect non-dopaminergic or cortical–limbic influences. Key limitations included the small sample size, which reduced statistical power, and reliance on self-reported exposure. Specifically, although exposure status was based on patient report, this was corroborated where possible using VA records, reducing—but not eliminating—misclassification risk. Furthermore, the lack of biochemical confirmation and a male-predominant cohort limited generalizability. Confirmation of distinct clinical profiles in exposed patients could improve diagnostic classification and prognostic counseling. With the 50th anniversary of the Vietnam War recently passing, the window to study this cohort is closing. Further research is essential to provide personalized care and inform policies protecting service members from environmental neurotoxins. If these exploratory findings are confirmed in larger cohorts, differential cognitive and patient-reported motor profiles may inform VA disability evaluations, prognostic counseling, and stratification in AO-exposed Veterans.

## Data Availability

All data produced in the present study are available upon reasonable request to the authors

